# Potential Antenatal Care-Mediated Benefits of Delivering Maternal Immunization in Five Low- and Middle-Income Countries: A Modeling Analysis

**DOI:** 10.64898/2026.03.03.26346908

**Authors:** Boshen Jiao, Isabelle Iversen, Ryoko Sato, Fentabil Getnet, Meseret Zelalem, Yohannes Lakew Tefera, Richmond Owusu, Josephine Gakii Gatua, Clint Pecenka, Sadaf Khan, Ranju Baral, Margaret E. Kruk, Catherine Arsenault, Stéphane Verguet

## Abstract

**Background:** Maternal immunization (MI) can prevent major infectious diseases in mothers and children by boosting the immunity of pregnant women. Antenatal care (ANC) delivery platforms could be leveraged to effectively provide MI. Adding MIs into ANC could potentially enhance ANC services, positively influencing both maternal and infant health outcomes and yielding broader benefits. We model these potential ANC-mediated health benefits in five low- and middle-income countries: Ethiopia, Ghana, Kenya, Pakistan, and South Africa.

**Methods:** We first developed a conceptual framework delineating pathways through which MI-ANC could enhance ANC utilization and quality, leading to improved care-seeking for facility delivery, postnatal care, and major childhood vaccinations (e.g., measles, diphtheria-pertussis-tetanus [DPT] third dose), as well as decreased infant mortality. Using a decision-analytic model informed by Demographic and Health Survey data, we simulated the potential benefits of MI-ANC delivery across socioeconomic groups at varying hypothetical MI coverage levels.

**Results:** MI-ANC integration would be associated with improvements in maternal and child health outcomes across all countries studied, mediated through enhanced engagement with ANC services. Under a scenario of full MI-ANC coverage, for example, infant mortality in Ethiopia’s poorest quintile was projected to decline from approximately 60 to 50 deaths per 1,000 live births. In-facility delivery rates were estimated to increase from 11% to 35%, postnatal care utilization from 4% to 11%, measles-containing vaccine coverage from 43% to 71%, and DPT3 immunization from 36% to 63%. These improvements would vary substantially by country and socioeconomic group, with the largest gains observed in populations with lower baseline ANC utilization.

**Conclusions:** Integrating MIs into ANC services has the potential to yield ANC-mediated health benefits, particularly in settings with low baseline ANC utilization. These findings can help inform priority-setting, support the design of targeted pilot programs, and guide future empirical implementation research on the possible broader impacts of MI-ANC delivery.

## Introduction

The World Health Organization (WHO) recommends various vaccinations for pregnant women (e.g., tetanus toxoid, influenza, and COVID-19 vaccines) owing to their potential to significantly avert maternal and child morbidity and mortality (1–3). These maternal vaccinations provide a direct defense against major infectious diseases, contributing to reduced maternal and infant mortality and morbidity (1–4). Moreover, recent strides in vaccine development have yielded new preventative options, like the respiratory syncytial virus (RSV) vaccine, newly approved for pregnant women to prevent RSV-related respiratory conditions in young infants (5,6). Despite the existing recommendations and new options, notable disparities in the uptake of maternal immunizations remain, especially in low- and middle-income countries (LMICs) (7,8), in part due to financial and operational constraints, among others (7).

The role of antenatal care (ANC) services is pivotal in this context, serving as an ideal platform for delivering these essential maternal immunizations (MIs) (9). ANC’s holistic approach includes the monitoring of potential complications, management of infections, chronic conditions and other potential obstetric risk factors, education on prenatal and perinatal care, and the promotion of nutritional and immunization protocols (10). WHO currently recommends at least eight ANC contacts to maximize maternal health and mothers’ satisfaction and optimize neonatal outcomes (4). Actual attendance, however, often falling short of meeting even the four-visit standard set by the previous WHO Focused Antenatal Care Model (11). According to UNICEF estimates, only 66% of pregnant women receive at least four ANC visits worldwide (10). Observations suggest that adequate and high-quality ANC utilization is strongly associated with better child health outcomes (9). A prior study analyzing data from 69 LMICs revealed that having at least four ANC visits and at least one encounter with a skilled provider was associated with an approximately 1.5% reduction in the probability of infant mortality (9). Beyond direct child health benefits, sufficient and high-quality ANC utilization is also associated with a higher uptake of other health services, such as facility deliveries (12,13), postnatal care (14,15), and adherence to infant immunization schedules (16,17), all contributing to overall health improvements.

These observed associations underscore the potential broader benefits of adding new MIs into ANC delivery platforms in LMICs. By doing so, it is anticipated that not only would the direct health benefits to mothers and children be enhanced, but large indirect gains in health outcomes could also occur, specifically by improving the use, range, and potential quality of ANC services (18,19). Such MI-ANC delivery might bolster the perceived value of these ANC services (18,19), thereby increasing trust among pregnant women, encouraging their participation and care-seeking, and eventually improving broader health outcomes. Despite this potential, empirical evidence quantifying such ANC-mediated health benefits in LMICs remains unexplored. More generally, the existing economic evaluation literature underscores the importance of considering the broader implications of health interventions on health systems beyond their direct impacts (20–22); yet, these considerations are often overlooked in priority-setting exercises.

In the absence of available empirical data, decision analytical modeling can be a valuable tool to integrate existing evidence from various sources and project the potential effects of new interventions (23), and evaluate the effect of not-yet-available technologies (24,25). In this study, we use a decision-analytic model to estimate the maximum potential ANC-mediated health benefits of introducing a hypothetical MI through ANC platforms in selected LMICs. This scenario-based analysis is intended to inform health policy design and guide future research and implementation priorities.

## Methods

### Overview

Our first step was to conceptualize the potential ANC-mediated benefits of adding a new MI into ANC services on health outcomes (Figure 1). Therein we proposed several interconnected pathways through which MI-ANC could be impactful. The first pathway postulated that the introduction of a new MI within ANC services may enhance both the utilization and quality of ANC services (26). Integrating a new vaccine into the package of ANC services may attract more women to seek ANC services, perceiving them as higher quality (26). The integration of a new vaccine could lead to better quality care through increased staff motivation and additional training, which ensures that healthcare providers are well-equipped to deliver comprehensible care (26,27). These augmentations are expected to lead to better child health outcomes, including perhaps a reduction in infant mortality rates (28), which constitutes the second pathway. Concurrently, the third pathway suggests that improvements in ANC may also positively affect the utilization of non-ANC health services, such as in-facility deliveries and postnatal care (28,29). Although we recognize the possibility of additional broader health system benefits, such as workforce strengthening, as well as improvements in maternal mental health resulting from MI-ANC, these are not explicitly modeled in our study due to a lack of available data.

**Figure 1.**
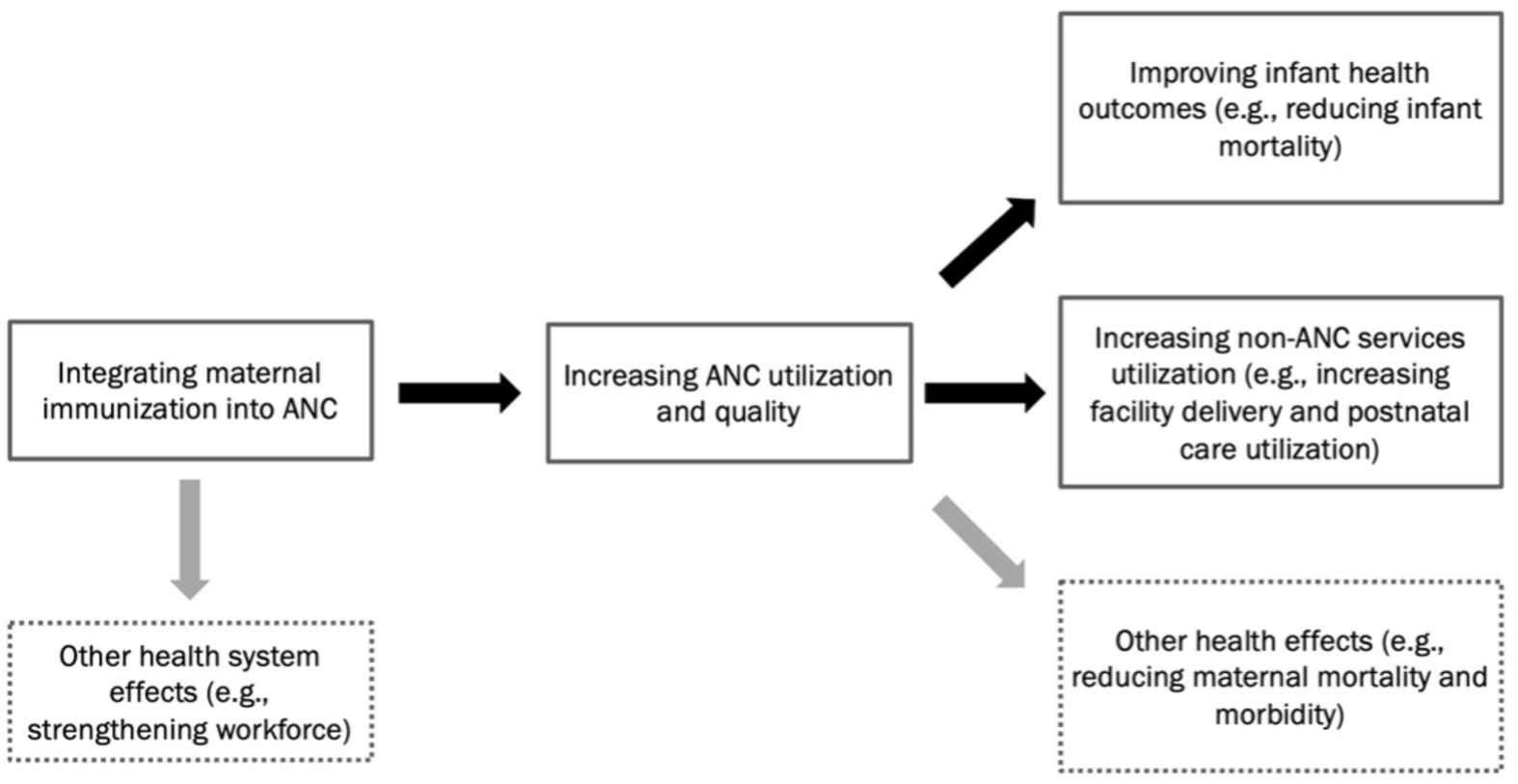
Conceptual framework for modeling the potential antenatal care (ANC)-mediated effects of adding maternal immunization into ANC). Note: Grey arrows indicate dimensions not modeled in our analysis, due to lack of empirical data.

To quantitatively assess these pathways, we constructed a decision analytic model that simulates the expected ANC-mediated health benefits across a spectrum of coverage rates for the hypothetical, novel, to-be-introduced, MI. We focused our analysis on five LMICs—Ethiopia, Ghana, Kenya, Pakistan, and South Africa—selected for their diverse geographic, socioeconomic, and epidemiological characteristics. Their gross national income (GNI) per capita spans from 1,000 USD to 6,800 USD (30). The WHO universal health coverage index in these countries varies between 45% and 71% (31), and the mortality rate for children under five years of age ranges from 33 to 63 per 1,000 live births (32). The model primarily evaluates the ANC-mediated effect of MI-ANC on the following outcomes: 1) infant mortality, defined as the probability of a child dying before her/his first birthday; 2) the effect on non-ANC service utilization, including in-facility delivery rates and postnatal care (e.g., check-ups in the first two months of life); and 3) child immunization coverage, that is coverage of diphtheria-pertussis-tetanus third dose (DPT3) and first dose of measles-containing vaccine (MCV1). The analysis adopts a per-pregnancy time horizon spanning pregnancy through early infancy (up to the first year of life), consistent with DHS-based measurement of the assessed outcomes.

### Decision analytic model

Decision-analytic modeling is a structured approach used to estimate health outcomes under alternative scenarios by integrating data from multiple sources and applying assumptions about intervention effects (23). This method enables the evaluation of potential impacts where empirical evidence is limited or unavailable.

The structure of our decision analytic model is depicted in Figure 2. Within this model, we stratified pregnant women into the following two cohorts to reflect real-world variability in immunization coverage: 1) those who received the new MI via ANC and 2) those who did not. For women not receiving the new MI through ANC, ANC service utilization and quality were presumed to remain consistent with current levels—some achieving what is considered adequate care and others not. Adequate ANC was defined as a minimum of four visits that encompassed essential service components, including blood pressure measurement, blood sample collection, and urine testing. Therefore, in the absence of the new MI, these women’s health outcomes would remain the same (that is, maintaining status quo).

**Figure 2.**
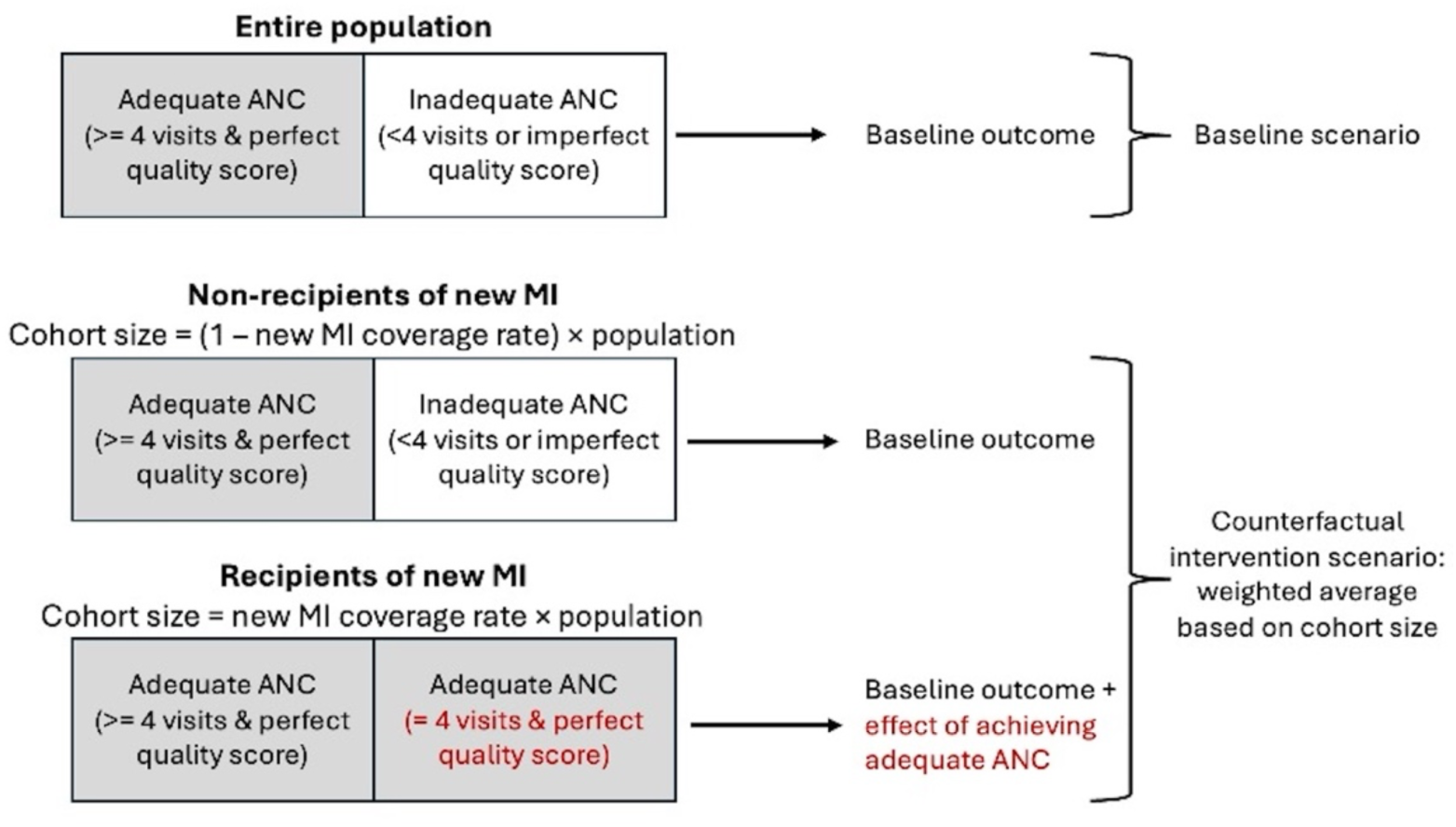
Schematic describing the decision analytic model for capturing the potential antenatal care (ANC)-mediated health effects of adding maternal immunization (MI) into ANC. Note: This model compares expected outcomes under two scenarios: the baseline scenario and the counterfactual intervention scenario. In the baseline scenario, outcomes reflect the current distribution of ANC utilization and quality in the population. In the counterfactual intervention scenario, we assume that women who would otherwise receive inadequate ANC achieve adequate ANC—defined as four visits with perfect quality—if they receive the new MI through ANC. Women who do not receive the new MI are assumed to maintain their baseline ANC status and associated outcomes. Thus, recipients of the new MI benefit from improved outcomes due to achieving adequate ANC, while non-recipients experience no change. Baseline outcomes are summarized in Table 1, and the estimated effects of achieving adequate ANC are provided in Table 2.

**Table 1.** Input parameters for the baseline outcomes across wealth quintiles, in Ethiopia, Ghana, Kenya, Pakistan, and South Africa.

| Country | Wealth quintile | Infant mortality (per 1,000 live births) | Facility delivery (%) | Postnatal care (%) | DPT3 immunization (%) | MCV1 immunization (%) |
| --- | --- | --- | --- | --- | --- | --- |
| Ethiopia | Poorest | 62 | 11 | 4 | 36 | 43 |
|  | Poorer | 55 | 19 | 5 | 50 | 50 |
|  | Middle | 60 | 22 | 8 | 51 | 54 |
|  | Richer | 75 | 27 | 10 | 63 | 59 |
|  | Richest | 54 | 69 | 17 | 76 | 74 |
| Ghana | Poorest | 55 | 46 | 73 | 87 | 87 |
|  | Poorer | 44 | 60 | 72 | 87 | 90 |
|  | Middle | 39 | 76 | 67 | 89 | 89 |
|  | Richer | 47 | 93 | 72 | 89 | 89 |
|  | Richest | 51 | 96 | 83 | 92 | 93 |
| Kenya | Poorest | 40 | 30 | 56 | 83 | 76 |
|  | Poorer | 42 | 49 | 65 | 90 | 87 |
|  | Middle | 39 | 62 | 69 | 92 | 88 |
|  | Richer | 46 | 80 | 72 | 94 | 95 |
|  | Richest | 38 | 93 | 77 | 93 | 93 |
| Pakistan | Poorest | 76 | 42 | 36 | 51 | 47 |
|  | Poorer | 71 | 52 | 26 | 72 | 69 |
|  | Middle | 67 | 69 | 28 | 76 | 77 |
|  | Richer | 51 | 81 | 33 | 88 | 84 |
|  | Richest | 53 | 92 | 37 | 91 | 88 |
| South Africa | Poorest | 54 | 92 | 82 | 72 | 85 |
|  | Poorer | 44 | 95 | 80 | 55 | 85 |
|  | Middle | 46 | 98 | 83 | 68 | 88 |
|  | Richer | 27 | 97 | 85 | 68 | 87 |
|  | Richest | 40 | 99 | 81 | 62 | 88 |

DPT3 = diphtheria-tetanus-pertussis third dose; MCV1 = measles-containing-vaccine first dose.

**Table 2.** Input parameters for the effect size (standard error in parentheses) on various outcomes of achieving adequate antenatal care (ANC), across wealth quintiles, in Ethiopia, Ghana, Kenya, Pakistan, and South Africa.

| Country | Wealth quintile | Infant mortality (per 1,000 live births) | Facility delivery (%) | Postnatal care (%) | DPT3 immunization (%) | MCV1 immunization (%) |
| --- | --- | --- | --- | --- | --- | --- |
| Ethiopia | Poorest | -11.6 (1) | 25.4 (1.8) | 5.4 (1.0) | 26.8 (1.1) | 30.0 (1.6) |
|  | Poorer | -9.8 (0.9) | 35.3 (2.7) | 7.4 (1.2) | 24.2 (3.6) | 15.6 (1.6) |
|  | Middle | -10.4 (1.1) | 23.7 (2.0) | 7.7 (1.1) | 19.8 (1.2) | 14.4 (3.0) |
|  | Richer | -2.6 (2.4) | 19.0 (2.1) | 11.0 (1.6) | 13.4 (2.3) | 15.1 (2.5) |
|  | Richest | -4.6 (0.4) | 16.3 (0.9) | 3.9 (0.5) | 10.7 (1.6) | 7.4 (0.8) |
| Ghana | Poorest | -1.6 (0.3) | 4.1 (0.6) | 2.7 (0.7) | 2.4 (0.6) | 1.9 (0.6) |
|  | Poorer | -0.8 (0.3) | 3.8 (0.7) | 1.3 (0.6) | 2.6 (0.7) | 2.1 (0.7) |
|  | Middle | 0.3 (0.6) | 1.4 (0.4) | 1.1 (0.3) | 1.4 (0.5) | 1.0 (0.5) |
|  | Richer | -0.1 (0.1) | 0.7 (0.2) | 0.1 (0.1) | -0.1 (0.1) | 0.1 (0.1) |
|  | Richest | 0.0 (0.1) | 0.6 (0.3) | 0.0 (0.1) | 0.0 (0.0) | 0.0 (0.0) |
| Kenya | Poorest | -2.0 (0.2) | 7.9 (0.4) | 1.0 (0.5) | 2.8 (0.3) | 3.6 (0.3) |
|  | Poorer | -1.1 (0.1) | 6.0 (0.6) | -0.3 (0.6) | 0.9 (0.2) | 1.7 (0.6) |
|  | Middle | -1.0 (0.1) | 4.5 (0.4) | -1.1 (0.4) | 1.0 (0.2) | 1.5 (0.3) |
|  | Richer | -0.6 (0.1) | 2.5 (0.2) | 0.6 (0.3) | 0.4 (0.2) | 1.1 (0.1) |
|  | Richest | 0.0 (0.2) | 0.7 (0.1) | 0.0 (0.1) | 0.1 (0.1) | 0.3 (0.1) |
| South Africa | Poorest | -2.7 (1.5) | 17.7 (2.5) | 3.2 (1.5) | 10.1 (0.9) | 10.2 (0.9) |
|  | Poorer | -1.4 (1.1) | 25.6 (2.0) | 3.7 (1.8) | 5.3 (0.7) | 4.8 (0.6) |
|  | Middle | -1.8 (0.6) | 7.9 (1.7) | 2.5 (0.8) | 1.8 (2.3) | 6.4 (2.3) |
|  | Richer | -1.9 (0.4) | 7.2 (1.2) | 1.1 (0.6) | 2.6 (1.3) | 5.2 (1.0) |
|  | Richest | -0.4 (0.1) | 1.4 (0.5) | 0.7 (0.7) | 0.4 (0.2) | 0.9 (0.2) |
| Pakistan | Poorest | -0.7 (0.2) | 0.9 (0.4) | 0.9 (0.3) | 1.1 (0.4) | 0.5 (0.2) |
|  | Poorer | -0.6 (0.1) | 0.2 (0.2) | 2.4 (0.6) | 0.1 (0.3) | 0.6 (0.3) |
|  | Middle | -0.3 (0.1) | 0.1 (0.1) | 0.7 (0.2) | 0.1 (0.7) | 0.9 (0.4) |
|  | Richer | -0.3 (0.1) | 0.1 (0.1) | 0.2 (0.4) | 0.1 (0.7) | 0.3 (0.1) |
|  | Richest | -0.3 (0.1) | 0.0 (0.0) | 0.6 (0.3) | 0.7 (0.5) | 0.8 (0.7) |

DPT3 = diphtheria-tetanus-pertussis third dose; MCV1 = measles-containing-vaccine first dose.
Note: The effect size, shown with the standard error in parentheses, measures the change associated with achieving the recommended levels of ANC visits across five wealth quintiles and multiple countries. The change represents predicted differences in various outcomes under two scenarios: 1) a scenario where all women achieve the recommended levels of ANC utilization and quality, and 2) a baseline scenario reflecting the current state of ANC in each country.

In contrast, for those receiving the new MI through ANC, we assumed that women who would have otherwise received adequate ANC would maintain their current level of care. For women who would not have received adequate ANC, the model does not simulate a behavioral process through which additional ANC visits are accumulated. Instead, receipt of the new MI through ANC is treated as a marker of successful engagement with an enhanced ANC platform, and these women are assumed to experience the average health outcomes associated with achieving adequate ANC, defined as at least four visits with complete service content. This assumption is intended to capture potential improvements in ANC utilization and quality attributable to MI-ANC integration, rather than to imply a literal increase in the number of visits for each individual.

While this assumption reflects a favorable scenario, it is grounded in both qualitative insights and quantitative evidence. Expert consensus and prior literature suggest that embedding a new MI delivery within ANC platforms has the potential to enhance service provision and increase patient uptake (27,33–35). Moreover, prior studies of similar maternal vaccines, such as tetanus toxoid, have shown strong associations with improved ANC utilization and adherence to care quality standards (36–40). These findings lend empirical support to the plausibility of our modeled assumptions. Nonetheless, it should be noted that this reflects an upper-bound scenario. Its purpose is to inform the potential maximum ANC-mediated health benefits of MI-ANC integration and to help prioritize future research and policy efforts in settings where such gains may be most achievable.

### Model parameters

Baseline health outcomes for each country were sourced from the latest available Demographic and Health Surveys (DHS) (41). The ANC-mediated effects of achieving adequate ANC services on non-ANC health service outcomes in the population were derived from a recently published parent study (29). This study analyzed DHS data from 2010 to 2022 across 47 LMICs. It employed logistic regression to estimate outcomes for two different scenarios: 1) the baseline, representing existing levels of ANC and 2) a counterfactual intervention scenario. The intervention scenario would elevate ANC visits to a minimum of four visits for individuals with fewer than four visits in the baseline, and improved ANC quality to the perfect level for those with imperfect baseline ANC quality. The difference in outcome estimates between these scenarios would represent the average effect of receiving adequate ANC at the population level. The analysis was conducted with stratification by both country and wealth quintile. A summary of this parent study is available in supplementary webappendix 1. For infant mortality, the parent study did not provide estimates; hence, we applied the same econometric approach to derive these figures. All parameters used in the model are detailed in Tables 1 and 2. Access to DHS data was obtained through the DHS program’s formal data request process. All analyses were conducted in accordance with the program’s terms of use.

### Analysis

We investigated the ANC-mediated effects of the hypothetical MI-ANC delivery for coverage rates ranging from 0% (indicating no joint MI-ANC delivery, that is the status quo) to 100% (indicating full coverage). The analysis was conducted in a stratified manner, considering both wealth quintiles within each country and differences across the countries themselves. To project expected estimates and capture uncertainty, we employed a probabilistic model. We assumed that the baseline rates of health outcomes followed a binomial distribution and that the effect sizes of achieving adequate ANC followed a normal distribution. We ran n=1,000 iterations of Monte Carlo simulations, the expected outcomes of which were then computed as the average of these simulations, with 95% simulation-based uncertainty ranges determined by the 2.5^th^ and 97.5^th^ percentiles of the simulated estimates.

All statistical analyses were performed using RStudio (Version 2023.09.1+494). Patients and members of the public were not involved in the design, conduct, reporting, or dissemination plans of this research.

### Patient and Public Involvement

Patients and members of the public were not involved in the development of the research question, outcome measures, study design, recruitment, conduct, or dissemination plans of this modeling study. As this analysis was based on secondary data from publicly available sources, no patient advisers were involved. There was no direct patient burden assessed.

## Results

The results (Figure 3) indicate that MI-ANC delivery would likely positively affect various health outcomes for mothers and children across the five countries examined in the study and their respective wealth quintiles. These health outcomes include reductions in infant mortality and increases in in-facility delivery rates, postnatal care rates, and child immunization coverage for both MCV1 and DPT3.

**Figure 3.**
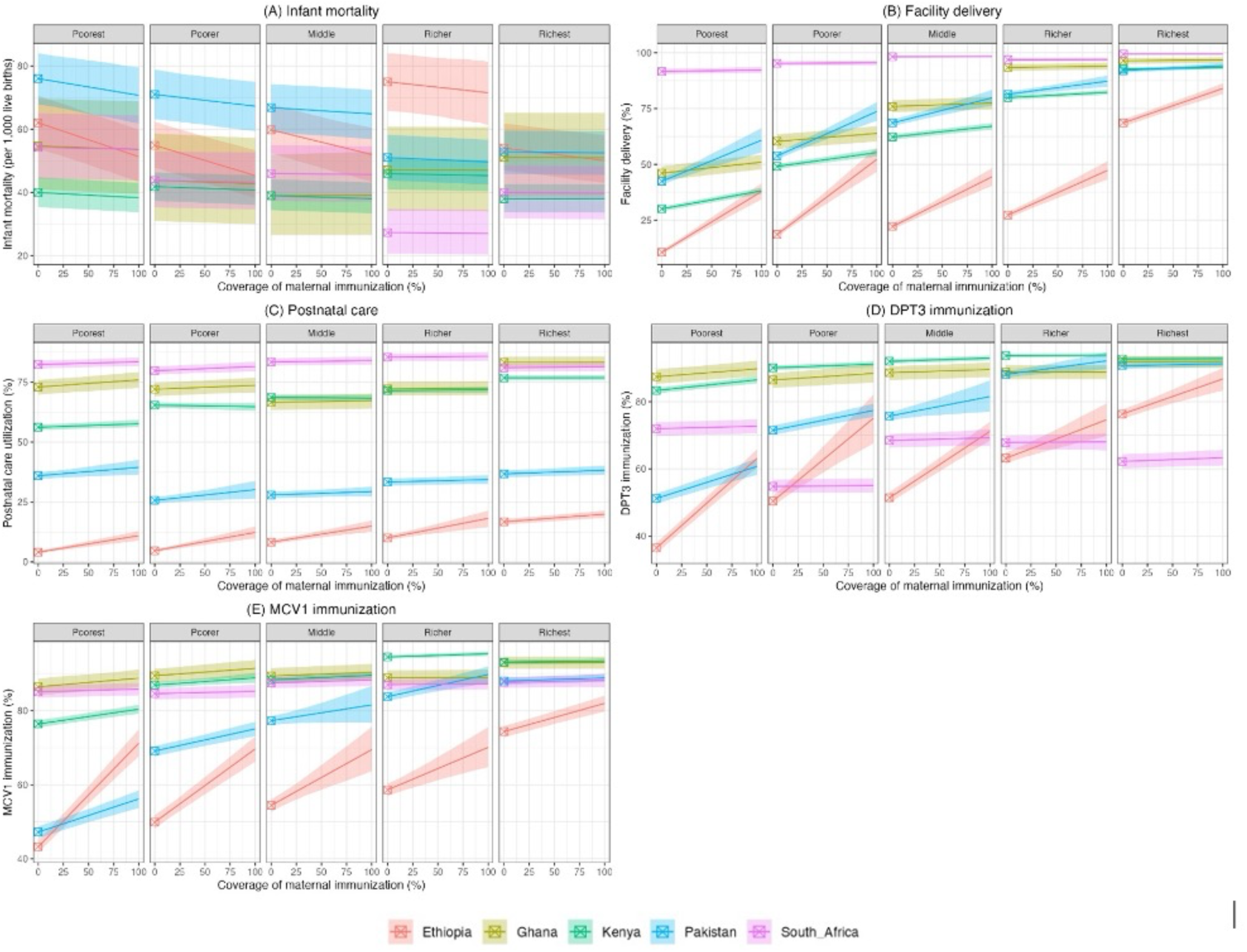
Potential antenatal care (ANC)-mediated effects of integrating maternal immunization on infant mortality (A), facility delivery (B), postnatal care utilization (C), diphtheria-tetanus-pertussis third dose (DPT3) vaccination (D), and first dose of measles-containing-vaccine (MCV1), by wealth quintile, in Ethiopia, Ghana, Kenya, Pakistan, and South Africa.

Notably, the extent of the ANC-mediated benefits would correlate with the coverage rate of the MI introduced; yet, important variations would occur depending on the country and across wealth quintiles. Overall, all countries in the study experienced a positive effect when MI would be added to the ANC platform, with larger effects estimated in the countries with lower baseline coverage. Regardless of wealth quintile, Ethiopia and Pakistan would exhibit the most substantial ANC-mediated effects of MI-ANC integration on reducing infant mortality and on increasing in-facility delivery rates, postnatal care rates, and child immunization coverage for both MCV1 and DPT3. Conversely, the effect was less pronounced in South Africa, Ghana, and Kenya.

Focusing on specific wealth quintiles, the model predicts that with 100% MI-ANC coverage in Ethiopia’s poorest quintile, infant mortality would decrease from approximately 60 deaths per 1,000 live births to 50 per 1,000. Additionally, in-facility delivery rates would be expected to rise from 11% to 35%, postnatal care rates from 4% to 11%, MCV1 immunization from 43% to 71%, and DPT3 immunization from 36% to 63%. Furthermore, in Ethiopia’s wealthiest quintile, we would see a reduction in infant mortality from 54 to 50 per 1,000 live births, with more modest increases in facility delivery (from 69% to 84%), postnatal care (from 17% to 20%), DPT3 immunization (from 76% to 87%), and MCV1 immunization (from 74% to 82%). Similarly, in Pakistan, the model predicted a relatively larger effect of the intervention on all these health outcomes for the poorest quintile.

In contrast, South Africa’s poorest quintile, even with 100% coverage, would see minimal changes. Infant mortality would remain at approximately 50 deaths per 1,000 live births, facility delivery rates would stay around 92%, postnatal care utilization would increase slightly from 82% to 83%, and immunization rates for DPT3 and MCV1 would slightly increase from 72% to 73% and 85% to 86%, respectively. Similarly, in Ghana and Kenya, the potential ANC-mediated effects estimated would be minimal, even for the poorest quintile.

## Discussion

Our decision modeling analysis conducted in Ethiopia, Ghana, Kenya, Pakistan, and South Africa suggests that MI-ANC delivery could have positive ANC-mediated benefits on maternal and child health outcomes and non-ANC services. Our findings indicate possible increases in in-facility delivery rates, postnatal care checks, and improved coverage of DPT3 and MCV1 immunizations. The magnitude of those benefits would vary largely across countries and wealth quintiles, being more pronounced in countries like Ethiopia and Pakistan with lower baseline coverage, especially among the lower wealth quintiles of these populations.

The variations observed could be attributed to differences in baseline ANC utilization and quality levels, which vary substantially from country to country and across socioeconomic groups. For instance, in Ethiopia, where baseline ANC levels were the lowest, the implementation of adequate ANC through MI could result in large improvements (42). In contrast, South Africa, with higher baseline ANC coverage levels (42), would only see modest gains. Furthermore, poorer individuals generally experience lower levels of ANC utilization and quality (43), suggesting that added services via MI-ANC would disproportionately benefit the poorest in all countries.

Variations in baseline coverage of non-ANC services between countries (e.g., differences in facility delivery rates) also contribute to the heterogeneity in the estimated ANC-mediated benefits. In countries with lower baseline facility delivery rates, such as Ethiopia, the potential for improvement through MI-ANC would likely be higher (43). Since richer women are already more likely to deliver at health facilities (44), strengthening ANC via MI-ANC would predominantly benefit countries and groups with lower utilization, possibly enhancing equity both within and across countries.

While the direct effects of MIs on infectious disease prevention are well established (45), our study is one of the first to explore the hypothetical benefits of adding such immunizations into ANC on a selected spectrum of maternal and child health outcomes via enhancing ANC engagement. There is an increasing acknowledgment of the broader value of health interventions beyond their primary intent, including the potential to strengthen health systems at large (20–22). Such potential benefits, though crucial, are frequently overlooked in conventional economic evaluations and priority-setting exercises (21,22). Our study lays out a framework to capture the potential ANC-mediated benefits of weaving a new intervention into existing health systems, offering a broader impact assessment approach. By focusing on MI as a means to strengthen ANC services, our study contributes new insights to this area of research. Indeed, it is important to recognize that health system strengthening effects could extend beyond the specific outcomes evaluated here. For instance, adding novel immunizations into ANC could also enhance the capabilities of the healthcare workforce within ANC services (18,19). This dual advantage–direct benefits and systemic improvements–should be a key consideration in shaping health policies for mothers and children.

The results of this study hold implications for LMICs like Ethiopia. Previously, the tetanus toxoid vaccine was the sole immunization offered to pregnant women within Ethiopia’s routine immunization program, and has now been replaced by the tetanus-diphtheria (Td) vaccine since 2019 (46). Our analysis underscores the potential benefits of MI-ANC to further broader health outcomes within the Ethiopian health system. With essential maternal and child health services like skilled delivery and postnatal care remaining below desired levels in Ethiopia (43), incorporating the ANC-mediated effects of MI-ANC into planning can elevate the importance of MI within ANC services, alongside standard practices such as blood pressure monitoring, hemoglobin testing, and iron supplementation. Additionally, with 15% of pregnant women in Ethiopia having Group B *Streptococcus* (GBS) recto-vaginal colonization (47), merging MI efforts with child immunization programs could be pivotal in the country by facilitating completion of immunization schedules, including the five-dose Td series. Effectively implemented, MI-ANC delivery could play a crucial role from childbirth onward, enhancing overall maternal and child health outcomes.

In Ghana, MI is currently part of ANC, with about 74% of women receiving sufficient tetanus toxoid injections to protect their offspring against neonatal tetanus in 2022 (48). This high coverage may be attributed to MI-ANC integration as an estimated 98% of pregnant women had at least one ANC visit and 88% had four or more visits (48). According to the Ghana National Reproductive Health Policy and Standards, MI is a core activity of ANC services (49); therefore, the gains from the integrated MI-ANC for tetanus toxoid could be extended to new vaccines such as RSV and GBS. The high prevalence of GBS (especially among rural communities) (50) makes the integration of the new vaccine into ANC more imperative. Since November 2022, Ghana Health Service has been working to integrate infectious diseases screening (including GBS) into ANC in five districts with a possible national scale-up after piloting (51). These experiences alongside MI-ANC integration for tetanus toxoid can be leveraged for all the new vaccines that pregnant women will need to protect their newborns.

Similarly, Kenya has been recognized for successfully eliminating maternal and neonatal tetanus, with vaccine coverage exceeding 80% in 2018 (42). The introduction of free maternal healthcare has facilitated tetanus toxoid MI implementation; however, the minimal effects of MI in ANC discussed in this paper may be attributed to the already high baseline coverage in Kenya. Consequently, the unmet need for vaccination might be low, resulting in only marginal benefits from additional interventions. It is important to note that certain individuals consistently missed by these efforts due to various unidentified factors could benefit from alternative strategies. Although the effect estimated here is limited, it does not imply that further improvement in MI coverage in Kenya is not possible. Addressing this will require a deeper understanding of the populations not being reached and targeting them effectively by addressing specific barriers.

While improvements in the uptake of maternal health services have been documented in Pakistan, the country continues to lag on key maternal and newborn health indicators. For example, in 2017-2018, the neonatal mortality rate was 42 deaths per 1,000 live births and the maternal mortality ratio was 186 deaths per 100,000 live births (52). Contributors include poor uptake, quality, and continuity of ANC. While uptake of ANC was near universal, only about half of women received four or more ANC visits, resulting in major missed opportunities (52). A DHS analysis indicated that only a minority of women in Pakistan receive quality ANC services (53). Similarly, the Expanded Programme on Immunization has continued to face challenges due to geopolitical, logistic, capacity, and sociocultural issues resulting in low coverage among target populations (54). This is also reflected in tetanus toxoid vaccination coverage among pregnant women, with 63% reporting receipt of two doses (per the latest DHS) (52). As of December 2023, Pakistan is one of only 11 countries that have failed to meet the targets for global maternal and neonatal tetanus elimination (55). With the inherent weaknesses in reach and coverage of immunization and ANC services, the program has undertaken supplemental immunization activities to reach pregnant women. Introducing new vaccines “based on evidence and epidemiological data” is one key objective stated under the 2022 National Immunization Policy (56). Integrated delivery of current and new maternal vaccines provides an opportunity to address the gaps in providing both ANC and vaccines and to improve the range of services offered via ANC. Results from this analysis indicate supplemental, ANC-mediated benefits with implications of substantive improvement in key indicators for Pakistan. This, in turn, could help the country make progress on meeting its commitments under the Sustainable Development Goals while achieving key priorities and objectives under national policies (57).

This study presents a scenario-based modeling analysis of delivering MI through ANC as a strategy for health system strengthening. A key strength of this work is the novel application of a decision-analytic framework to estimate the potential ANC-mediated benefits of MI-ANC integration—an area underexplored in the literature. By leveraging nationally representative data from five LMICs, the model captures variation across diverse health system contexts and socioeconomic groups. These insights offer valuable guidance for future empirical research and policy and planning. For instance, future implementation studies could assess the real-world impact of integrating new MIs, such as RSV or GBS, into ANC platforms. Additionally, policymakers may consider piloting MI-ANC delivery in underserved populations, where the potential health gains and system-wide improvements are likely to be substantial. Such targeted interventions could yield critical empirical evidence to inform scale-up decisions and support more equitable maternal and child health outcomes.

Nevertheless, our study has several limitations. First, we assumed that women receiving the new MI would experience improved engagement with ANC as a result of MI-ANC integration. In practice, this assumption may not hold universally, as structural, geographic, and financial barriers may limit women’s ability to attend additional ANC visits, even when new services such as maternal vaccination are offered. As such, our findings should be interpreted as representing an upper-bound scenario, intended to illustrate the maximum potential ANC-mediated benefits of integrating MI into ANC rather than to predict real-world outcomes. This limitation highlights the need for future empirical research to quantify the extent to which offering MI through ANC translates into increased ANC attendance and engagement across different settings and populations. Second, our model simulated a random distribution of MI in populations, leading to a linear relationship between increased coverage and improved outcomes. This assumption may not hold for women already accessing adequate ANC who are more likely to receive the immunization, potentially resulting in a non-linear relationship. Third, the impacts of the integration strategy might extend beyond the outcomes examined in this study, including broader effects on the health system (e.g., workforce enhancement), maternal risk behaviors and comorbidities, child morbidity, the long-term productivity effects of improved child health outcomes, and the downstream effects of non-ANC health services (e.g., potential child mortality reduction due to increased measles vaccination rate). These omissions could lead to an underestimation of the full benefits of MI-ANC delivery. Fourth, the model does not explicitly incorporate several real-world implementation factors that may influence the feasibility and effectiveness of MI-ANC integration. These include vaccine hesitancy, redistribution of staff workload, and potential negative spillovers, such as overburdened health workers having less time to deliver routine ANC components (33,34). Such constraints are particularly salient in fragile or resource-limited health systems, where introducing additional services without adequate support could inadvertently compromise care quality. Consequently, realizing the potential benefits of MI-ANC integration in practice may require complementary system-level investments, including strengthening workforce capacity, engaging local stakeholders, identifying operational bottlenecks, and adopting context-specific implementation strategies to ensure that MI delivery enhances, rather than strains, existing ANC services. Fifth, the effect estimates for achieving adequate ANC services are subject to potential confounders that may remain unobserved despite thorough adjustments for known confounders (58). For instance, the quality of follow-up care that some women receive at home could account for the observed minimal impact on non-ANC service utilization, such as facility deliveries.

## Conclusions

Our decision analytic model underscores the potential of adding novel MIs into ANC services. The findings advocate for a decision-making approach that considers both direct and indirect ANC-mediated benefits of new MIs in maternal and child health policy and planning. Future research should focus on empirically testing these effects in real-world settings with pilot implementations, particularly targeting the most underserved groups in countries with the greatest ANC disparities.

## Data Availability

All data produced in the present study are available upon reasonable request to the authors.

## Webappendix 1

### Summary of the parent study

The parent study (1) analyzed data from 78 Demographic and Health Surveys from 2010 to 2022 across 47 low- and middle-income countries. It employed multilevel logistic regression to explore the impact of achieving antenatal care (ANC) on a series of outcomes such as facility delivery, postnatal care, and childhood immunizations. The model included predictors like the number of ANC visits and its square term, an ANC quality score, and fixed effects for country and wealth quintile, while adjusting for confounders. Interaction terms between ANC variables, country, and wealth quintile were incorporated to address heterogeneity in ANC impacts across different contexts. The study predicted outcomes under two scenarios using the fitted regression model: 1) a baseline scenario reflecting the current pattern of ANC, observed in the DHS data, and 2) a counterfactual intervention scenario. In the counterfactual scenario, the intervention hypothesized increasing ANC visits to four for individuals with fewer than four visits in the baseline and enhancing ANC quality scores to one for individuals with lower baseline scores. The difference in projected outcomes between these scenarios was interpreted as the potential impact of attaining adequate ANC. This analysis was stratified by wealth quintile and country. The resulting effect size estimates used in this modeling study are summarized in detail in Table 2.

